# Tau-Clinical Mismatch Identifies Individuals with Co-Pathology and Predicts Clinical Trajectory

**DOI:** 10.1101/2025.07.25.25332195

**Authors:** Christopher A. Brown, Nidhi S. Mundada, Katheryn A.Q. Cousins, Niyousha Sadeghpour, Xueying Lyu, Emily McGrew, Magdalena Korecka, Alice Chen-Plotkin, Long Xie, Laura E.M. Wisse, John A. Detre, Corey T. McMillan, Edward B. Lee, Ilya M. Nasrallah, Sandhitsu R. Das, Dawn Mechanic-Hamilton, Paul A. Yushkevich, Leslie M. Shaw, Alzheimer’s Disease Neuroimaging Initiative, David A. Wolk

## Abstract

**Importance:** The heterogeneous course of Alzheimer’s disease makes it difficult to predict individuals’ cognitive trajectories, which is particularly important in the era of disease modifying therapy. Identifying individuals more likely to have co-pathology and differing disease courses using clinically practical tools remains a critical gap.

**Objective:** To evaluate tau-clinical mismatch for identifying resilient and vulnerable individuals and compare levels of co-pathology and clinical trajectories between groups.

**Design, Setting, and Participants:** Participants were selected from the Alzheimer’s Disease Neuroimaging Initiative (ADNI, inclusion from 2005-2024), Penn Alzheimer’s Disease Research Center Cohort (Penn-ADRC, inclusion from 2002-2025), and Penn Anti-amyloid Therapy Monitoring (Penn-ATM) cohort (inclusion from 2024-2025). All participants were amyloid-β positive, had clinical assessment, and measures of Tau-PET or plasma p-tau217 available.

**Exposures:** Clinical assessment (CDR-SB) and tau burden (tau-PET or p-tau_217_) for mismatch group classification.

**Main Outcomes and Measures:** Cross-sectional measures of neurodegeneration (medial temporal lobe volume and thickness, cortical thickness, TAR DNA-binding protein 43 [TDP-43] imaging signature), α-synuclein cerebrospinal fluid seed-amplification assay, longitudinal CDR-SB

**Results:** 365 ADNI Tau-PET participants (ages 55-93, 52.6% women) and 524 ADNI p-tau_217_ participants (ages 56-95, 49.0% women) were used to generate tau-clinical mismatch models with 55.6-57.1% classified as canonical (CDR-SB ∼ Tau), 23.7-24.7% as resilient (CDR-SB < Tau), and 19.3-19.7% as vulnerable (CDR-SB > Tau). Groups showed diverging clinical courses with earlier cognitive impairment seen in vulnerable groups and later impairment in resilient groups. Vulnerable groups showed higher frequencies of co-pathology, with TDP-43 neurodegeneration patterns and α-synuclein positivity. Similar findings were seen when applying these models to an independent dataset of 244 individuals (54-92 age, 57.0% women) in Penn-ADRC. Finally, these models were applied to a cohort receiving anti-amyloid therapy to show the utility of this method for predicting individual cognitive trajectories during therapy.

**Conclusion and Relevance:** Tau-clinical mismatch identifies individuals more likely to harbor co-pathology and have diverging clinical trajectories. Plasma-based models produced similar results to Tau-PET models and could be replicated in independent datasets. These models provide an important tool that can be implemented in clinical practice to provide improved individualized prognosis and, potentially, monitoring of response to disease-modifying therapy.

## Introduction

There is significant heterogeneity in the clinical progression of Alzheimer’s disease (AD)^1,2^. While some of this heterogeneity is explained by level of tau burden and neurodegeneration, large inter-individual differences in disease trajectory exist even at similar levels of AD pathology^3^. One known driver of heterogeneity in clinical trajectory is the presence or absence of co-pathologies, which are present in up to 65-70% of individuals with AD at autopsy^4^. The presence of these co-pathologies, as well as inter-individual differences in brain resilience and cognitive reserve, make it difficult to assess the degree to which AD pathology is contributing to clinical impairment at the patient level, as well as to predict the likely clinical trajectory^3,5,6^. This represents a critical gap in the era of disease-modifying therapy, where treatment decisions and monitoring would greatly benefit from the ability to provide individualized guidance on disease trajectory, the relative contribution of different pathologies, and, in turn, likely therapeutic benefit^2^.

One potential method for assessing both co-pathology and resilience is to examine the mismatch between the degree of tau burden and clinical impairment. Prior work has demonstrated that mismatch between tau and neurodegeneration using structural MRI or ^18^F-FDG-PET can identify individuals with TAR-DNA binding protein-43 (TDP-43) and alpha-synuclein (αSyn) co-pathology^7–10^. Further, individuals exhibiting greater neurodegeneration than expected for a given level of tau show faster clinical decline, while those with less neurodegeneration than expected show slower decline^7,8^. The recently revised criteria for the diagnosis and staging of AD by the Alzheimer’s Association workgroup and recent attempts to operationalize these criteria have further highlighted the concept of tau-clinical (T-C) mismatch as a potentially valuable tool to identify these resilient and vulnerable groups^11,12^.

Therefore, we hypothesized that T-C mismatch will identify individuals on different clinical trajectories, reflecting underlying differences in co-pathology burden and cognitive reserve. To test this hypothesis, we used both T_1_ and T_2_ biomarkers of tau pathology, as defined by the new criteria^11^, with clinical assessment in the Alzheimer’s Disease Neuroimaging Initiative (ADNI) dataset to develop and evaluate models of T-C mismatch. Unlike the recently revised criteria which defines tau and clinical stages in a discrete manner, here we considered them using continuous measures. We assessed the presence of αSyn and likely TDP-43 co-pathology, as well as longitudinal clinical trajectories in ADNI prior to applying models to our Penn Alzheimer’s Disease Research Center Clinical Core Cohort (Penn-ADRC) for replication. Finally, we illustrated the potential clinical utility of these markers for individualized monitoring during anti-amyloid therapy in our Penn ADRC Anti-amyloid Therapy Monitoring (Penn-ATM) cohort.

## Methods

### Participants

Individuals were selected from the ADNI dataset based on availability of: 1) Positive amyloid status in the “UCBERKELEY_AMY_6MM” csv, 2) at least one Tau-PET or Fujirebio Lumipulse plasma p-tau_217_, and 3) at least one Clinical Dementia Rating Scale Sum of Boxes (CDR-SB) score. All participants also had an MRI available and a subset had available cerebrospinal fluid Amprion αSyn seed amplification assay (SAA) and longitudinal CDR-SB. Data were collected October 2005-December 2024. All participants provided informed consent as detailed in the ADNI protocol. For Penn-ADRC, participants were selected based on availability of: 1) Fujirebio Lumipulse p-tau_217_/Aβ_42_ ratio > 0.0055 (amyloid positive cutoff)^13^, and 2) at least one CDR-SB score. All participants also had an MRI available and a subset had longitudinal CDR-SB. Data were collected May 2002-January 2025. Finally, Penn-ATM participants were included based on availability of 1) Fujirebio Lumipulse p-tau_217_, 2) MMSE, and 3) Dementia Severity Rating Scale (DSRS)^14^. Data were collected May 2024-January 2025. All Penn participants provided informed consent under protocols approved by the University of Pennsylvania Institutional Review Board.

### Assessment of Tau Pathology

^18^F-flortaucipir tau-PET acquisition and plasma collection were performed as previously described and are detailed in the eMethods^15^. For assessment of global tau burden in the tau-PET dataset, partial volume corrected (PVC) Standardized Uptake Value Ratio (SUVR) were used to calculate Global Tau-MaX, which provides a region-agnostic assessment of global tau magnitude and extent with values ranging from 0 to 100^15^. Fujirebio Lumipulse p-tau_217_ was used as a measure of tau burden in the ADNI plasma, Penn-ADRC and Penn-ATM datasets.

### Clinical Assessment

For both ADNI and Penn-ADRC, CDR-SB were obtained for the time-point closest to Tau-PET or p-tau_217_ for cross-sectional analyses and all scores were included for longitudinal analyses. For the Penn-ATM dataset, the closest MMSE and DSRS prior to p-tau_217_ collection was used for analyses.

### Assessment of Neurodegeneration and Co-Pathology Signatures

T1-weighted and fluid-attenuated inversion recovery magnetic resonance imaging (MRI) sequences were used to assess multiple structural measures (Supplement 1 for acquisition parameters). For both ADNI and Penn-ADRC, T1-weighted images were processed using the ANTs Cortical Thickness pipeline and T1-Automated Segmentation of Hippocampal Subfields (ASHS) and Cortical Reconstruction for ASHS (CRASHS) pipelines^16–20^. We also calculated a TDP-43-related MRI signature consisting of the ratio of entorhinal cortex (ERC) thickness to parahippocampal cortex (PHC) thickness, which is reduced in autopsy-proven TDP-43+ individuals^21,22^. Finally, we evaluated α-synuclein seed amplification assay in cerebrospinal fluid as measure of αSyn co-pathology in ADNI^23^. Additional details for these methods are provided in the eMethods in Supplement 1.

### Statistical Analyses

All analyses were performed in R v.4.4.1 with additional visualizations performed using ParaView and SurfIce. Prior to statistical analyses, outliers identified as > 5 SDs from the mean were excluded. We first used linear regression to examine the association of Tau-MaX with p-tau_217_, while controlling for centiloids to confirm the relationship between Tau-MaX and p-tau_217_. We then used separate linear regressions to model the relationship between CDR-SB and 1) Tau-MaX or 2) p-tau_217_, while controlling for age, sex, and years of education. Standardized residuals (SR) were extracted for each participant to classify individuals into canonical (-0.6 ≤ SR ≤ 0.6), vulnerable (SR > 0.6), and resilient (SR < -0.6). While this cutoff is based on prior work^10^, sensitivity analyses repeated cross-sectional analyses with mismatch residual as a continuous measure (see Supplement 1). We then compared Tau-MaX and p-tau_217_ mismatch in a subset of participants with both measures available within 2 years of each other. Next, we examined differences between mismatch groups in brain structure and measures of co-pathology while controlling for age, sex, education, and global tau burden (Tau-MaX or p-tau_217_). For MTL surface-based analyses, threshold-free cluster enhancement FWE correction was used with *p_corr_* < .05 considered significant. For whole-brain ROI analyses, FDR correction was used with *p*_corr_ < .05 considered significant. Linear models were used for continuous outcome measures and logistic regression was used for dichotomous outcomes.

For longitudinal analyses, we first used sampled iterative local approximation (SILA) to determine an individual’s estimated tau onset age (ETOA) using either Tau-PET or plasma p-tau_217_ (see Supplement 1 for details)^24^. We then examined CDR-SB trajectory using linear mixed-models with a Mismatch Group × time from ETOA (using b-spline with 3 degrees of freedom) as the predictor of interest, age, sex, education, and baseline tau burden as covariates, and a random effect of participant. Next, we used SILA to estimate cognitive impairment onset age (ECOA) defined as CDR-SB ≥ 4 with time from ETOA as the age input; Survival analyses examined differences in time without cognitive impairment (from ETOA to ECOA) between mismatch groups using Cox-proportional hazards models.

We then applied the ADNI p-tau_217_ mismatch model to Penn-ADRC to generate predicted CDR-SB and regressed against actual CDR-SB to extract residuals for mismatch classification based on the ADNI model. We repeated classification within Penn-ADRC by repeating the same steps performed in ADNI and compared this classification with the classification from the applied ADNI model. Next, we evaluated cross-sectional differences in brain structure, longitudinal differences in CDR-SB trajectory and survival to cognitive impairment (ECOA) between mismatch groups as described above. Finally, we sought to illustrate the potential implementation of these models in clinical practice using baseline data from the Penn ATM cohort. We used a cross-walk from MMSE and DSRS to CDR-SB based on historical Penn ADRC data (see Supplement 1) and p-tau_217_ to classify individuals into mismatch groups, determine ETOA, and then predict their expected longitudinal trajectory over the course of treatment.

## Results

### Mismatch Classification and Concordance Between T_1_ and T_2_ Biomarkers

365 Aβ+ individuals with Tau-PET and 524 Aβ+ individuals with plasma p-tau_217_ were included in analyses. Tau-MaX and P-tau_217_ were strongly associated (*β*_std_ = 0.63 [0.54, 0.72], *t*(255) = 13.6, *p* < .001), even after controlling for centiloids (eFigure 1). There was a positive association between both Tau-MaX (*β*_std_ = 0.53 [0.44, 0.61], *t*(360) = 11.7, *p* < .001) and p-tau_217_ (*β*_std_ = 0.47 [0.39, 0.54], *t*(319) = 12.4, *p* < .001) with CDR-SB (Figure 1A). For Tau-PET, 203 (55.6%) were classified as canonical, 90 (24.7%) as resilient, and 72 (19.7%) as vulnerable, and for p-tau_217_, 299 (57.1%) were classified as canonical, 124 (23.7%) as resilient, and 101 (19.3%) as vulnerable. In individuals with both biomarkers available, 182/258 (71%) had concordant classification and only 1/258 (0.4%) had vulnerable-resilient discordance (Figure 1A).

**Figure 1.**
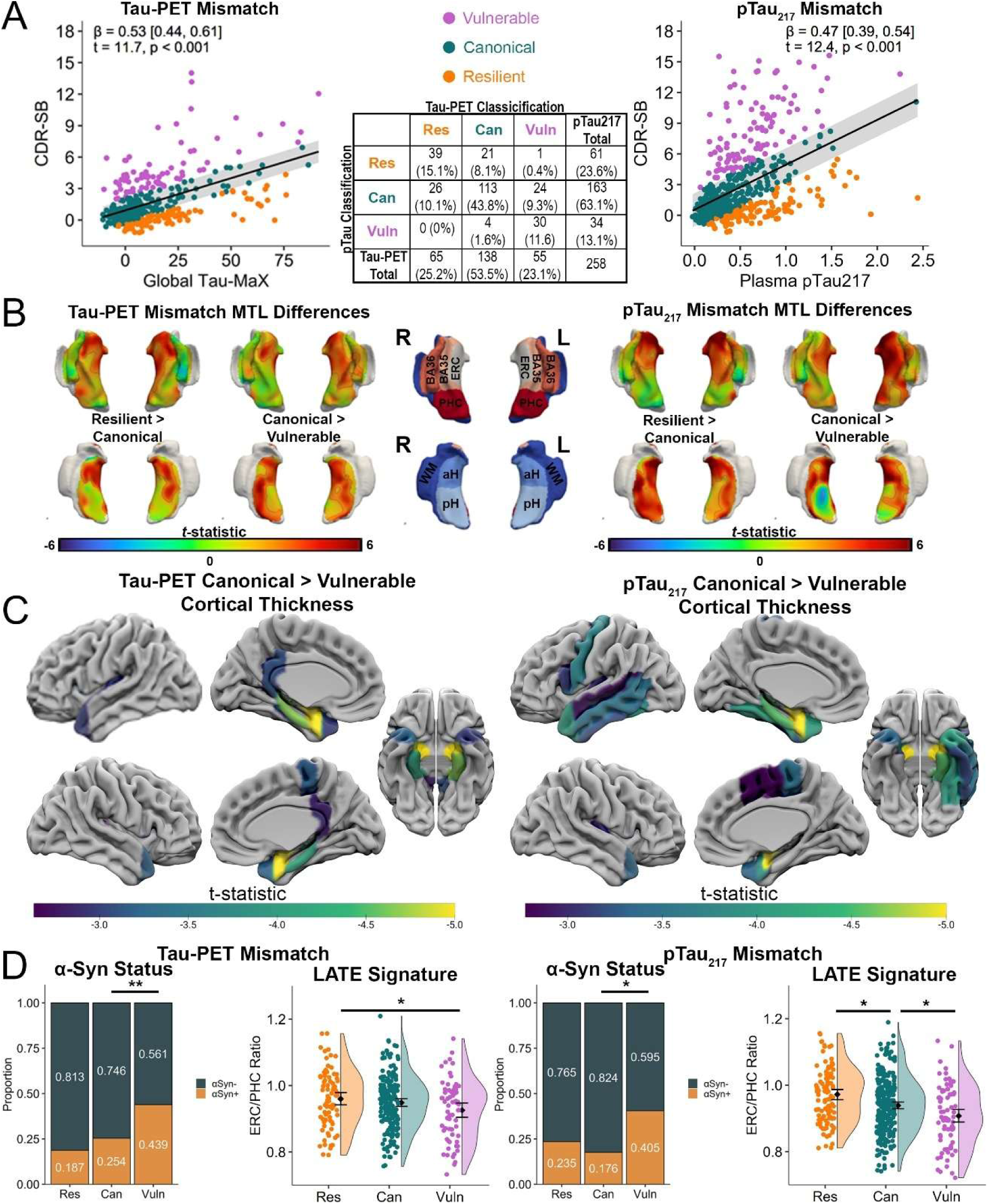
Cross-Sectional Differences in Atrophy and Co-pathology between ADNI Mismatch Groups. **A:** Association between CDR-SB and Tau-MaX (left) and p-tau_217_ (right) with mismatch classification shown by color scale. Gray shading represents Standardized Residual (SR) = 0.6. The confusion matrix for mismatch classification within a subset of participants with both Tau-MaX and p-tau_217_ available is shown in the middle. **B:** Differences in MTL thickness between mismatch groups for Tau-MaX (left) and p-tau_217_ (right) with color-scale representing t-stat for group comparison and black lines outlining clusters surviving TFCE *p*_FWE_ < .05. Vulnerable groups show primarily decreased thickness in anterior MTL regions along with posterior hippocampus, while resilient groups greater thickness in a similar regional pattern. aH (anterior hippocampus), pH (posterior hippocampus), ERC (entorhinal cortex), BA (Brodmann area), PHC (parahippocampal cortex). **C:** Comparison of cortical thickness between Tau-MaX (left) and p-tau_217_ (right) mismatch groups across whole-brain cortical ROIs with t-stat for group comparison shown and only regions surviving *p*_FDR_ < .05 shown. **D:** αSynuclein status and LATE-MRI signature ERC/PHC ratio compared between Tau-MaX (left) and p-tau_217_ (right) mismatch groups; \**p* < .05, \*\**p* < .005.

### Cross-sectional Comparison Between Mismatch Groups

Demographics and summary measures for mismatch groups are shown in Table 1. Canonical participants tended have younger age, higher female prevalence, and less AD pathology compared to both resilient and vulnerable groups. Vulnerable participants had greater clinical impairment than resilient and canonical groups. For cortical thickness, the vulnerable group had lower thickness predominantly in anterior MTL regions (Figure 1B) and temporopolar and fronto-opercular regions (Figure 1C) compared to the canonical group for both Tau-PET and plasma datasets. The resilient group showed higher thickness in similar anterior MTL regions (Figure 1B), although no differences were seen when examining across the whole brain after FDR-correction. For the Tau-PET dataset, vulnerable participants had marginally lower ERC/PHC ratio (*β_std_* = -0.23 [-0.51, 0.04], *t*(329) = -1.66, *p* = .099) compared to the canonical group (Figure 1D). For the p-tau_217_ dataset, the vulnerable group had a lower ERC/PHC ratio (*β_std_* = -0.28 [-0.53, -0.02], *t*(448) = -2.15, *p* = .032) compared to the canonical group, while the resilient group had higher ERC/PHC (*β_std_* = 0.43 [0.19, 0.67], *t*(448) = 3.51, *p* < .001) ratio compared to the canonical group. Finally, for both datasets, the vulnerable group had a higher rate of αSyn+ compared to the canonical group (Figure 1D, Tau-MaX: OR = 3.08 [1.52, 6.26], *p* = .002; p-tau_217_: OR = 1.86 [1.04, 3.33], *p* = .036).

**Table 1.** Group demographics and summary measures.

|  | ADNI Tau-PET Mismatch |  |  | ADNI pTau<sub>217</sub> Mismatch |  |  | Penn-ADRC Mismatch |  |  |
| --- | --- | --- | --- | --- | --- | --- | --- | --- | --- |
|  | Can<br>(n=203) | Res<br>(n=90) | Vuln<br>(n=72) | Can<br>(n=299) | Res<br>(n=124) | Vuln<br>(n=101) | Can<br>(n=182) | Res<br>(n=27) | Vuln<br>(n=35) |
| Age | 73.6<br>(7.22) | 78.5<br>(7.97) | 76.5<br>(8.27) | 75.6<br>(7.63) | 79.5<br>(7.87) | 78.5<br>(7.91) | 73.3<br>(6.89) | 75.2<br>(6.95) | 74.6<br>(5.98) |
| Sex |  |  |  |  |  |  |  |  |  |
| F | 115<br>(56.7%) | 42<br>(46.7%) | 35<br>(48.6%) | 162<br>(54.2%) | 49<br>(39.5%) | 45<br>(44.6%) | 111<br>(61%) | 12<br>(44.4%) | 16<br>(45.7%) |
| M | 88<br>(43.3%) | 48<br>(53.3%) | 37<br>(51.4%) | 137<br>(45.8%) | 75<br>(60.5%) | 56<br>(55.4%) | 71<br>(39%) | 15<br>(55.6%) | 19<br>(54.3%) |
| Education | 16.6<br>(2.24) | 15.6<br>(2.47) | 15.6<br>(2.58) | 16.5<br>(2.38) | 15.6<br>(2.83) | 15.8<br>(2.6) | 16.4<br>(2.57) | 14.9<br>(2.89) | 16.3<br>(2.92) |
| Diagnosis |  |  |  |  |  |  |  |  |  |
| CU | 93<br>(45.8%) | 52<br>(57.8%) | 0<br>(0%) | 123<br>(41.1%) | 61<br>(49.2%) | 0<br>(0%) | 84<br>(46.2%) | 17<br>(63.0%) | 0<br>(0%) |
| MCI | 89<br>(43.8%) | 33<br>(36.6%) | 18<br>(25%) | 117<br>(39.1%) | 52<br>(41.9%) | 3<br>(3%) | 64<br>(35.2%) | 9<br>(33.3%) | 6<br>(17.1%) |
| Dem | 21<br>(10.3%) | 5<br>(5.6%) | 54<br>(75%) | 59<br>(19.7%) | 11<br>(8.9%) | 98<br>(97%) | 34<br>(18.7%) | 1<br>(3.7%) | 29<br>(82.9%) |
| ApoE ε4 |  |  |  |  |  |  |  |  |  |
| 0 | 66<br>(39.5%) | 41<br>(50.6%) | 19<br>(30.2%) | 108<br>(41.2%) | 54<br>(47%) | 28<br>(28.9%) | -- | -- | -- |
| 1 | 81<br>(48.5%) | 29<br>(35.8%) | 31<br>(49.2%) | 127<br>(48.5%) | 44<br>(38.3%) | 53<br>(54.6%) | -- | -- | -- |
| 2 | 20<br>(12%) | 11<br>(13.6%) | 13<br>(20.6%) | 27<br>(10.3%) | 17<br>(14.8%) | 16<br>(16.5%) | -- | -- | -- |
| MMSE | 27.9<br>(2.46) | 27.7<br>(2.29) | 23<br>(4.48) | 26.9<br>(3.58) | 27.2<br>(2.98) | 18.7<br>(6.3) | -- | -- | -- |
| Centiloids | 68.2<br>(38.8) | 79.7<br>(37.5) | 89.5<br>(34.6) | 70.2<br>(38.3) | 87.2<br>(36.6) | 93.8<br>(38.1) | -- | -- | -- |
| Tau-MaX | 7.99<br>(16.0) | 20.1<br>(23.9) | 17.9<br>(22.3) | 11.4<sub>163</sub><br>(19.0) | 20.1<sub>61</sub><br>(25.4) | 24.3<sub>34</sub><br>(27.1) | -- | -- | -- |
| p-tau<sub>217</sub> | 0.39<sub>138</sub><br>(0.34) | 0.59<sub>65</sub><br>(0.43) | 0.64<sub>55</sub><br>(0.40) | 0.37<br>(0.30) | 0.70<br>(0.43) | 0.65<br>(0.37) | 0.41<br>(0.31) | 0.85<br>(0.54) | 0.59<br>(0.37) |
| CDR-SB | 0.98<br>(1.32) | 0.56<br>(0.82) | 5.21<br>(2.69) | 1.57<br>(1.86) | 0.83<br>(1.15) | 8.59<br>(3.31) | 1.26<br>(1.41) | 0.57<br>(1.05) | 7.39<br>(3.45) |
| αSyn |  |  |  |  |  |  |  |  |  |
| Neg | 123<br>(81.5%) | 50<br>(74.6%) | 32<br>(56.1%) | 179<br>(76.5%) | 75<br>(82.4%) | 47<br>(59.5%) | -- | -- | -- |
| Pos | 28<br>(18.5%) | 17<br>(25.4%) | 25<br>(43.9%) | 55<br>(23.5%) | 16<br>(17.6%) | 32<br>(40.5%) | -- | -- | -- |
| ERC/PHC | 0.96<sub>186</sub><br>(0.09) | 0.95<sub>82</sub><br>(0.09) | 0.92<sub>68</sub><br>(0.10) | 0.94<sub>264</sub><br>(0.09) | 0.96<sub>108</sub><br>(0.08) | 0.90<sub>80</sub><br>(0.09) | 0.98<sub>156</sub><br>(0.09) | 0.98<sub>22</sub><br>(0.07) | 0.91<sub>31</sub><br>(0.13) |

Mean (S.D) for continuous measures and count (%) for categorical variables. Subscripts denote the number of participants with data available for continuous measures when incomplete. Can: Canonical, Res: Resilient: Vuln: Vulnerable; F: Female, M: Male, CU: Cognitively Unimpaired, MCI: Mild Cognitive Impairment, Dem: Dementia αSyn: αSynuclein, Neg: Negative, Pos: Positive, ERC/PHC: Entorhinal cortex thickness/parahippocampal cortex thickness

### Longitudinal Clinical Trajectories

For linear mixed effect models in both the Tau-PET and plasma datasets, the vulnerable group showed faster clinical progression (Tau-MaX: *F*_3,1537_ = 22.3, *p* < .001; p-tau_217_: *F*_3,2815_ = 75.4, *p* < .001) and the resilient group showed slower clinical progression (*F*_3,1529_ = 17.5, *p* < .001; *F*_3,2770_ = 86.1, *p* < .001) compared to the canonical group (Figure 2A). After using SILA to determine ECOA, survival analyses showed vulnerable participants had higher risk for developing cognitive impairment over time (Tau-MaX: HR = 2.54 [1.93, 3.35], *p* < .001; p-tau_217_: HR = 3.04 [2.41, 3.86], *p* < .001) and resilient participants had lower risk for developing cognitive impairment over time (HR = 0.43 [0.33, 0.57], *p* = .004; HR = 0.21 [0.16, 0.27], *p* < .001) compared to canonical participants (Figure 2B).

**Figure 2.**
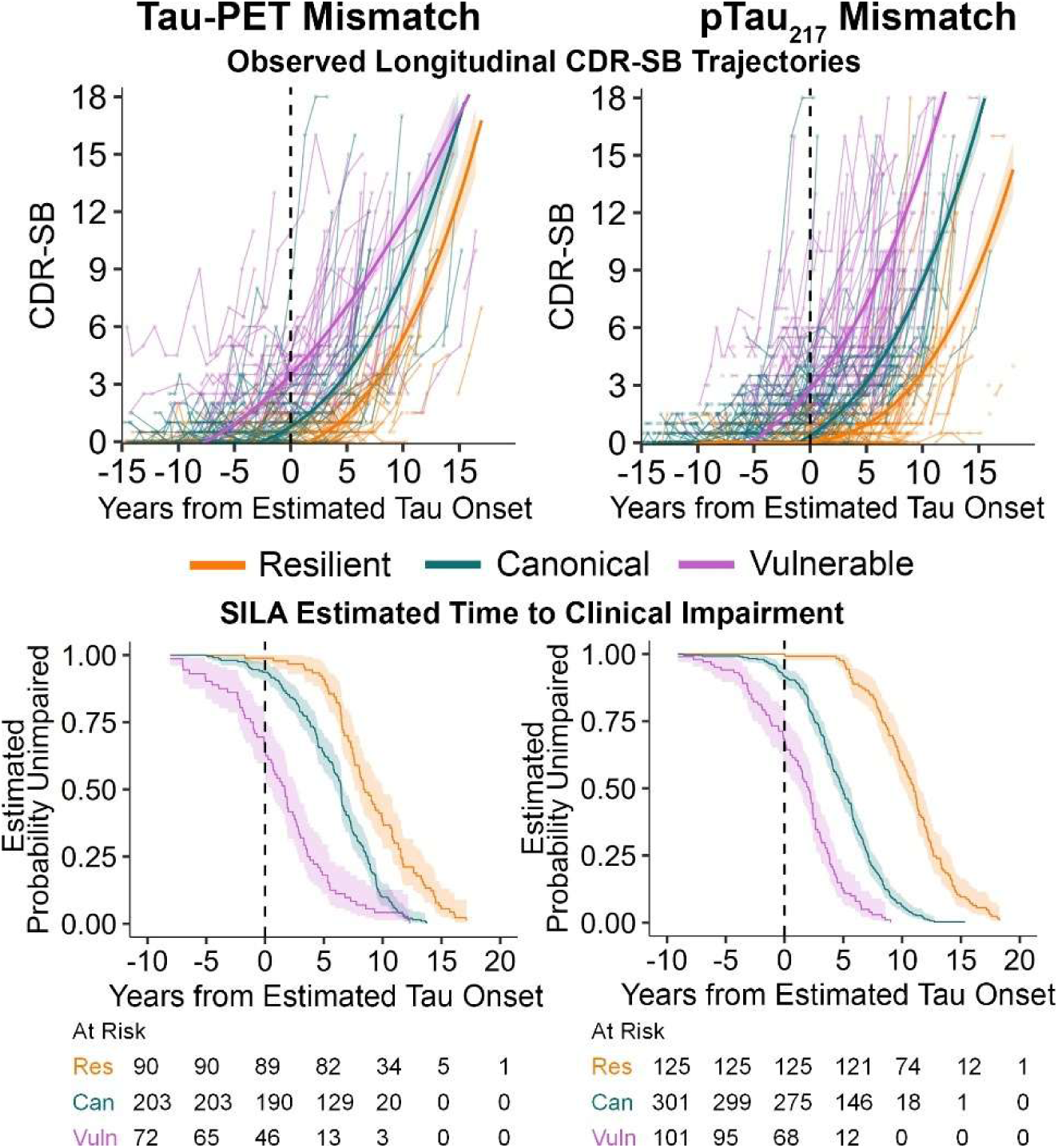
Diverging Longitudinal Clinical Trajectories between Mismatch Groups. **Top:** Longitudinal CDR-SB plotted against time from Tau positivity for Tau-MaX (left) and p-tau_217_ (right) mismatch groups. Each point represents individual time points with thin lines connecting time points from the same participant. Thick lines represent the b-spline best-fit for each mismatch group (color) with ribbon showing 95% confidence interval of fit. **Bottom:** Time surviving without clinically significant impairment (defined as CDR-SB < 4) based on years of tau positivity for Tau-MaX (left) and p-tau_217_ (right) mismatch groups. Lines represent the survival curve for each mismatch group (color) with ribbon showing the 95% confidence interval of fit.

### Replication in Penn-ADRC

In 244 Aβ+ individuals in Penn-ADRC, we found 87.7% concordance in mismatch classification when applying the ADNI p-tau_217_ mismatch model compared to regenerating the model within Penn-ADRC (Figure 3A). We found similar group differences to those seen in ADNI with vulnerable participants showing reduced thickness predominantly in anterior MTL and temporopolar regions (Figure 3B). The ERC/PHC ratio was lower (*β_std_* = -0.57 [-0.95, -0.18], *t*(202) = -2.91, *p* = .004) in vulnerable compared to canonical participants (Figure 3C). Finally, vulnerable participants showed faster clinical decline (*F*_3,1260_ = 11.9, *p* < .001) and higher risk of cognitive impairment over time (HR = 3.07 [2.10, 4.50], *p* < .001), while resilient participants showed slower clinical decline (*F*_3,1200_ = 27.8, *p* < .001) and lower risk of cognitive impairment over time (HR = 0.28 [0.18, 0.43], *p* < .001) compared to canonical participants (Figure 3D).

**Figure 3.**
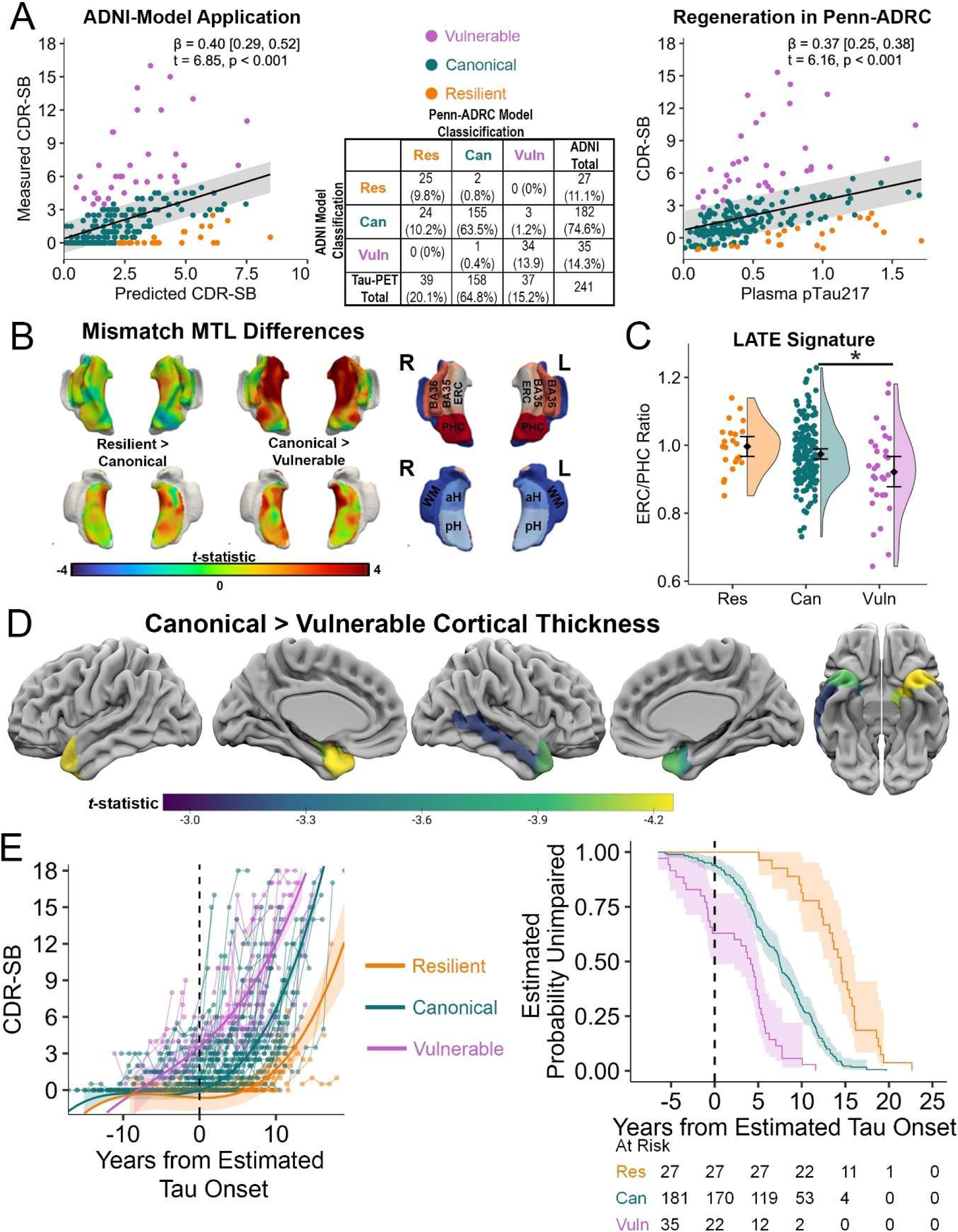
Replication in Penn ADRC Cohort. **A:** CDR-SB plotted against ADNI-model predicted CDR-SB (left) and p-tau_217_ within ABC (right) with ADNI-based mismatch classification shown by color scale for both. Gray shading represents Standardized Residual (SR) = 0.6. Confusion matrix for mismatch classification using the ADNI-derived and Penn ADRC-derived models are shown in the middle. **B:** Differences in MTL thickness between mismatch groups with color-scale representing the t-statistic for group comparison and black lines outlining clusters surviving TFCE *p*_FWE_ < .05. **C:** Comparison of the LATE MRI signature ERC/PHC thickness ratio between mismatch groups, *\*p* < .05. **D:** Comparison of cortical thickness between mismatch groups across whole-brain cortical ROIs with *t*-stat for group comparisons shown by color-scale and only regions surviving *p*_FDR_ < .05 shown. **E:** Longitudinal CDR-SB plotted against time from Tau positivity for mismatch groups (left). Each point represents individual time points with thin lines connecting time points from the same participant. Thick lines represent the b-spline best-fit for each mismatch group (color) with ribbon showing 95% confidence interval of fit. Time surviving without clinically significant impairment (defined as CDR-SB < 4) based on years of tau positivity for mismatch groups (right). Lines represent the survival curve for each mismatch group (color) with ribbon showing the 95% confidence interval of fit.

### Application to Penn-ATM Cohort

We then applied the ADNI p-tau_217_ model to the Penn-ATM cohort, where 71% of participants receiving anti-amyloid therapy were classified as canonical, 13% as resilient, and 16% as vulnerable (Figure 4A). Using the SILA and cognitive trajectory models described above, we predict the average expected change in CDR-SB over the 18-month treatment period, illustrating that expected change in CDR-SB differs depending on tau burden and mismatch classification (Figure 4B-D).

**Figure 4.**
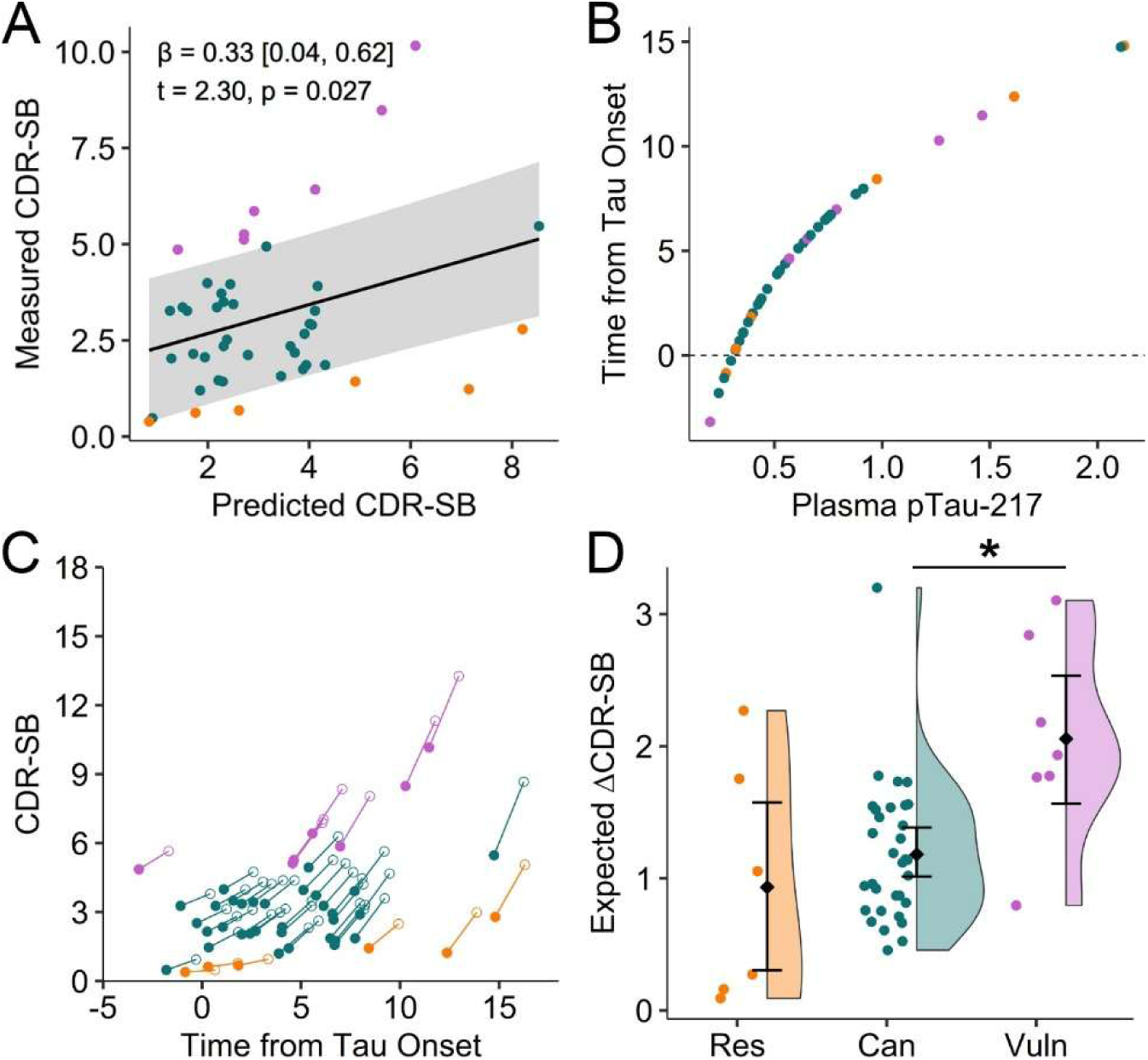
Application of mismatch models to Penn anti-amyloid therapy monitoring (ATM) cohort. **A**: Classification of individuals into mismatch groups (color) by comparison of measured CDR-SB against CDR-SB predicted by ADNI mismatch model. Gray ribbon represents the threshold of SR > 0.6 from the ADNI model. **B:** Calculation of time from tau-positivity using p-tau_217._ **C:** Predicting change in CDR-SB over treatment time using ADNI mismatch models. Closed circles are the actual CDR-SB measured at baseline and open circles are predicted CDR-SB after 18 months using longitudinal models generated from ADNI with color indicating mismatch group. **D:** Expected change in CDR-SB over 18 months of treatment by mismatch group shows significant differences based on mismatch classification (right). \**p* < .05. Res: Resilient, Can: Canonical, Vuln: Vulnerable

## Discussion

In this quantitative evaluation of T-C mismatch using T_1_ (p-tau_217_) and T_2_ (Tau-PET) markers of tau pathology, we found that individuals classified as cognitively vulnerable appear to exhibit higher levels of co-pathology and more rapid clinical decline even when accounting for level of AD pathology. These findings were consistent whether using a T_1_ or T_2_ marker of tau pathology and were replicated when applied to an independent dataset. Our results provide a model for assessing the likelihood of co-pathology and predicted disease course that could be particularly useful for monitoring disease progression and treatment response at the individual level, as highlighted by the application of this model to a cohort receiving anti-amyloid therapy. Further, stratification of patients based on the likelihood of co-pathology may provide insights into the relative efficacy of specific therapeutic interventions and ultimately influence clinical decisions for implementation.

Our findings were consistent with the hypothesis that individuals showing T-C vulnerability would be more likely to harbor co-pathology. This converges with prior work looking at disease progression in individuals with co-pathologies, studies using Tau-Neurodegeneration mismatch, and recent work operationalizing the revised AD diagnostic and staging criteria^8,10,12^. Similar to this work, we identified a pattern of reduced brain structure in anterior MTL, temporopolar, and fronto-opercular regions, which strongly overlaps with brain regions impacted in Limbic-predominant Age-related TDP-43 Encephalopathy (LATE)^22,25^. Consistent with this, we found that a structural imaging biomarker of LATE, the ERC/PHC ratio^21^, was lower in vulnerable groups. Moreover, we also found higher levels of αSyn co-pathology in the vulnerable groups within ADNI, which has also been reported previously using an analogous approach^12^.

In addition, our results supported the hypothesis that T-C mismatch would identify groups with diverging clinical trajectories. This, again, mirrors the literature examining clinical trajectory in the presence of co-pathology compared to pure AD^3,4^, as well as studies of Tau-Neurodegeneration mismatch^7,8^. However, T-C mismatch likely incorporates differential susceptibility to AD pathology and/or cognitive reserve, the latter of which may not be captured by approaches focusing only on biomarkers of neurodegeneration (e.g. brain structure), but likely plays an important role in clinical trajectory^5,26,27^. Nevertheless, the role of non-AD-related changes are evidenced by the fact that 33-37% of vulnerable individuals already show cognitive impairment by the time of tau positivity. In contrast, the canonical group does not reach this level until about 5 years after tau positivity, which is consistent with prior work examining progression to cognitive impairment in individuals who are either Tau-PET+ or p-tau_217_+^28^. In contrast, resilient individuals do not show similar rates of impairment until more than 10 years after tau positivity, which likely reflects a combination of brain reserve/resilience and cognitive reserve^8,27^.

We found consistent results using T_1_ or T_2_ markers to define T-C mismatch, which agrees with prior findings that p-tau_217_ is strongly correlated with Tau-PET burden in Aβ+ individuals and performs similarly to Tau-PET in predicting clinical progression^28^. The AD diagnostic and staging criteria operationalize biological staging with Tau-PET, whereas p-tau_217_, as a T_1_ marker is conceptualized as being more linked to amyloid status^11,12^. However, our results, suggest that p-tau_217_, beyond its sensitivity to amyloid status, may be able to provide similar information about tau load, or disease stage, with much greater clinical accessibility compared to Tau-PET. Future studies using blood-based T_2_ biomarkers may help to further improve T-C mismatch models, but currently these measures remain in research use-only and lack robust, fully automated implementations that would make them clinically feasible^11^.

To illustrate the generalizability and clinical use of these models, we applied them to an independent dataset and a clinical cohort receiving anti-amyloid therapy. Applying the mismatch model to a new cohort resulted in similar classification compared to repeating the analysis within the cohort and similar patterns of neurodegeneration were seen in Penn-ADRC and ADNI when comparing between groups. Of note, the Penn-ADRC cohort is likely enriched for individuals with LATE due to ongoing studies focused on LATE that co-enroll into the ADRC. Further, there were similar rates of cognitive decline and longitudinal impairment-free survival between mismatch groups across ADNI and Penn-ADRC, highlighting the generalizability of these models. This could be particularly useful for monitoring response to anti-amyloid therapy, as illustrated by application to the Penn-ATM cohort. Using our mismatch model, individuals receiving anti-amyloid therapy have considerable heterogeneity in expected change in CDR-SB over the 18-month treatment period, thus providing more individualized null models that could serve as a means to measure treatment response in individual patients^2,28^.

### Strengths and Limitations

The strengths of our study include a data-driven approach comparing T_1_ and T_2_ biomarkers to classifying T-C mismatch and validating our models in an independent dataset. A main limitation of these cohorts is that both are volunteer-based cohorts focused on studying Alzheimer’s disease with predominantly highly educated individuals. Of note, the Penn ADRC cohort has greater racial representation of the US population compared to ADNI, thus providing evidence that these models may generalize to a broader population. An additional limitation is the lack of longitudinal cognitive assessment in the ATM cohort to validate these models for treatment response, but future studies will be able to investigate this application further. Finally, we do not have autopsy-confirmed pathology, which would provide greater evidence that the structural changes observed in the vulnerable group reflect underlying TDP-43 pathology. Future work in autopsy cohorts with antemortem biomarkers and cognitive assessment should investigate this further.

### Conclusions

We developed T-C mismatch models that predict presence of co-pathology and diverging clinical trajectories using both T_1_ and T_2_ biomarkers of disease pathology within two independent cohorts. These models provide a clinically-applicable tool for predicting disease progression and, potentially, prediction of treatment response to anti-amyloid therapy in real-world settings.

## Supporting information

eMethods and Results

## Data Availability

All requests for raw and analyzed data from the ABC cohort will be reviewed by the Penn Neurodegenerative Data Sharing Committee (PNDSC) and shared for appropriate uses through a data sharing agreement (https://www.pennbindlab.com/data-sharing). Anonymized data from ABC will be shared upon request to the corresponding author by a qualified academic investigator for the purpose of replicating procedures and results in this article. Data are not publicly available due to privacy protections outlined in the participant informed consent. Documents related to study protocols, informed consent and other documentation can similarly be made available upon request. All ADNI data are shared without embargo through the LONI Image and Data Archive (https://ida.loni.usc.edu/), a secure research data repository. Interested scientists may obtain access to ADNI imaging, clinical, genomic, and biomarker data for the purposes of scientific investigation, teaching, or planning clinical research studies. Access is contingent on adherence to the ADNI Data Use Agreement and the publications' policies (https://adni.loni.usc.edu/data-samples/access-data/).

## Acknowledgements

This work was supported by grants from the National Institutes of Health (P30-AG072979, RF1-AG069474, R01-AG056014, R01-AG055005, R01-AG072796, R25-NS065745, P01-AG084497), Pennsylvania Department of Health (2019NF4100087335), Alzheimer’s Association and Fred Barbara Erb Foundation (AACSF-23-1152241), and Delaware Community Foundation. Data collection and sharing for the Alzheimer’s Disease Neuroimaging Initiative (ADNI) is funded by the National Institute on Aging (National Institutes of Health Grant U19 AG024904). The grantee organization is the Northern California Institute for Research and Education. In the past, ADNI has also received funding from the National Institute of Biomedical Imaging and Bioengineering, the Canadian Institutes of Health Research, and private sector contributions through the Foundation for the National Institutes of Health (FNIH) including generous contributions from the following: AbbVie; Alzheimer’s Association; Alzheimer’s Drug Discovery Foundation; Araclon Biotech; BioClinica, Inc.; Biogen; Bristol-Myers Squibb Company; CereSpir, Inc.; Cogstate; Eisai Inc.; Elan Pharmaceuticals, Inc.; Eli Lilly and Company; EuroImmun; F. Hoffmann-La Roche Ltd and its affiliated company Genentech, Inc.; Fujirebio; GE Healthcare; IXICO Ltd.; Janssen Alzheimer Immunotherapy Research & Development, LLC; Johnson & Johnson Pharmaceutical Research &Development LLC; Lumosity; Lundbeck; Merck & Co., Inc.; Meso Scale Diagnostics, LLC; NeuroRx Research; Neurotrack Technologies; Novartis Pharmaceuticals Corporation; Pfizer Inc.; Piramal Imaging; Servier; Takeda Pharmaceutical Company; and Transition Therapeutics.

## References

1. Devanarayan V, Ye Y, Charil A, et al. Predicting clinical progression trajectories of early Alzheimer’s disease patients. Alzheimers Dement. 2023;20(3):1725–1738. doi:10.1002/alz.13565

2. Duara R, Barker W. Heterogeneity in Alzheimer’s Disease Diagnosis and Progression Rates: Implications for Therapeutic Trials. Neurotherapeutics. 2022;19(1):8–25. doi:10.1007/s13311-022-01185-z

3. Boyle PA, Yu L, Leurgans SE, et al. Attributable risk of Alzheimer’s dementia attributed to age-related neuropathologies. Ann Neurol. 2019;85(1):114–124. doi:10.1002/ana.25380

4. Robinson JL, Xie SX, Baer DR, et al. Pathological combinations in neurodegenerative disease are heterogeneous and disease-associated. Brain. 2023;146(6):2557–2569. doi:10.1093/brain/awad059

5. Hoenig MC, Bischof GN, Hammes J, et al. Tau pathology and cognitive reserve in Alzheimer’s disease. Neurobiology of Aging. 2017;57(1):1–7. doi:10.1016/j.neurobiolaging.2017.05.004

6. Abner EL, Kryscio RJ, Schmitt FA, et al. Outcomes after diagnosis of mild cognitive impairment in a large autopsy series. Annals of Neurology. Published online February 2017. doi:10.1002/ana.24903

7. Das SR, Lyu X, Duong MT, et al. Tau-atrophy variability reveals phenotypic heterogeneity in Alzheimer’s disease. Ann Neurol. 2021;90(5):751–762. doi:10.1002/ana.26233

8. Lyu X, Duong MT, Xie L, et al. Tau-neurodegeneration mismatch reveals vulnerability and resilience to comorbidities in Alzheimer’s continuum. Alzheimers Dement. 2023;20(3):1586–1600. doi:10.1002/alz.13559

9. Duong MT, Das SR, Khandelwal P, et al. Hypometabolic mismatch with atrophy and tau pathology in mixed Alzheimer’s and Lewy body disease. Brain. 2024;148(5):1577–1587. doi:10.1093/brain/awae352

10. Duong MT, Das SR, Lyu X, et al. Dissociation of tau pathology and neuronal hypometabolism within the ATN framework of Alzheimer’s disease. Nat Commun. 2022;13(1):1495. doi:10.1038/s41467-022-28941-1

11. Jack CR, Andrews JS, Beach TG, et al. Revised criteria for diagnosis and staging of Alzheimer’s disease: Alzheimer’s Association Workgroup. Alzheimer’s & Dementia. 2024;20(8):5143–5169. doi:10.1002/ALZ.13859

12. Pichet Binette A, Smith R, Salvadó G, et al. Evaluation of the Revised Criteria for Biological and Clinical Staging of Alzheimer Disease. JAMA Neurology. Published online May 19, 2025. doi:10.1001/jamaneurol.2025.1100

13. Cousins KAQ, Korecka M, Wan Y, et al. Comparison of plasma p-tau217/Aβ42, p-tau217, and Aβ42/Aβ40 biomarkers by race to detect Alzheimer’s disease. Alzheimer’s & Dementia. doi:10.1002/alz.70469

14. Xie SX, Ewbank DC, Chittams J, Karlawish JHT, Arnold SE, Clark CM. Rate of Decline in Alzheimer Disease Measured by a Dementia Severity Rating Scale. Alzheimer Disease & Associated Disorders. 2009;23(3):268. doi:10.1097/WAD.0b013e318194a324

15. Brown CA, Das SR, Cousins KAQ, et al. Tau Burden is Best Captured by Magnitude and Extent: Tau-MaX as a Measure of Global Tau. Published online January 14, 2025. doi:10.1101/2025.01.13.25320488

16. Xie L, Wisse LEM, Pluta J, et al. Automated segmentation of medial temporal lobe subregions on in vivo T1-weighted MRI in early stages of Alzheimer’s disease. Human Brain Mapping. 2019;40(12):3431. doi:10.1002/HBM.24607

17. Tustison NJ, Holbrook AJ, Avants BB, et al. Longitudinal Mapping of Cortical Thickness Measurements: An Alzheimer’s Disease Neuroimaging Initiative-Based Evaluation Study. Journal of Alzheimer’s Disease. 2019;71(1):165–183. doi:10.3233/JAD-190283

18. Yushkevich PA, Ittyerah R, Li Y, et al. Morphometry of medial temporal lobe subregions using high-resolution T2-weighted MRI in ADNI3: Why, how, and what’s next? Alzheimer’s & Dementia. Published online 2024. doi:10.1002/ALZ.14161

19. Wuestefeld A, Pichet Binette A, van Westen D, et al. Medial temporal lobe atrophy patterns in early-versus late-onset amnestic Alzheimer’s disease. Alzheimers Res Ther. 2024;16(1):204. doi:10.1186/s13195-024-01571-z

20. Sadeghpour N, Lim SA, Wuestefeld A, et al. Developing an anatomically valid segmentation protocol for anterior regions of the medial temporal lobe for neurodegenerative diseases. bioRxiv. Published online February 13, 2025:2025.02.11.637506. doi:10.1101/2025.02.11.637506

21. de Flores R, Wisse LEM, Das SR, et al. Contribution of mixed pathology to medial temporal lobe atrophy in Alzheimer’s disease. Alzheimers Dement. 2020;16(6):843–852. doi:10.1002/alz.12079

22. Wolk DA, Nelson PT, Apostolova L, et al. Clinical criteria for limbic-predominant age-related TDP-43 encephalopathy. Alzheimer’s & Dementia. 2025;21(1):e14202. doi:10.1002/alz.14202

23. Tosun D, Hausle Z, Iwaki H, et al. A cross-sectional study of α-synuclein seed amplification assay in Alzheimer’s disease neuroimaging initiative: Prevalence and associations with Alzheimer’s disease biomarkers and cognitive function. Alzheimer’s & Dementia. 2024;20(8):5114–5131. doi:10.1002/alz.13858

24. Betthauser TJ, Bilgel M, Koscik RL, et al. Multi-method investigation of factors influencing amyloid onset and impairment in three cohorts. Brain. 2022;145(11):4065–4079. doi:10.1093/brain/awac213

25. Bejanin A, Murray ME, Martin P, et al. Antemortem volume loss mirrors TDP-43 staging in older adults with non-frontotemporal lobar degeneration. Brain. 2019;142(11):3621–3635. doi:10.1093/brain/awz277

26. Singh-Manoux A, Marmot M, Glymor M, Sabia S, Kivimaki M, Dugravot A. Does cognitive reserve shape cognitive decline? Annals of Neurology. 2011;70(2):296–304. doi:10.1038/jid.2014.371

27. Stern Y, Arenaza-Urquijo EM, Bartrés-Faz D, et al. Whitepaper: Defining and investigating cognitive reserve, brain reserve, and brain maintenance. Alzheimer’s & Dementia. Published online 2018:1–7. doi:10.1016/j.jmarsys.2011.03.015

28. Ossenkoppele R, Salvadó G, Janelidze S, et al. Plasma p-tau217 and tau-PET predict future cognitive decline among cognitively unimpaired individuals: implications for clinical trials. Nat Aging. 2025;5(5):883–896. doi:10.1038/s43587-025-00835-z

