## Supplementary material for "Tau-Clinical Mismatch Identifies Individuals with Co-Pathology and Predicts Clinical Trajectory": eMethods and Results

*ADNI Cohort*: The ADNI was launched in 2004 as a public-private partnership, led by Principal Investigator Michael W. Weiner, MD. The primary goal of ADNI has been to test whether serial magnetic resonance imaging (MRI), positron emission tomography (PET), other biological markers, and clinical and neuropsychological assessment can be combined to measure the progression of mild cognitive impairment (MCI) and early Alzheimer's disease (AD). For up-to-date information, see www.adni-info.org. All Aβ+ participants (defined by Amyloid PET using ADNI protocols) with at least one Tau-PET or Fujirebio Lumipulse p-$tau_{217}$ and at least one CDR-SB score were included.

*Penn ABC*: The Penn ABC dataset is an ongoing longitudinal observational study with annual evaluation using the Uniform Data Set and consensus diagnosis[1]. Participants were initially selected based on availability of plasma p-$tau_{217}$ and $A\beta_{42}$ measured on the Fujirebio Lumipulse platform with a Aβ+ cutoff of p-$tau_{217}$/$A\beta_{42}$ > 0.0055 selected based on previous data[2]. Aβ+ participants who also had at least one available CDR-SB score were included in analyses.

*Penn ATM*: The Penn ATM dataset is an ongoing longitudinal observational study of individuals with MCI or Dementia due to AD receiving anti-amyloid therapy at the University of Pennsylvania who provide their clinical data and assessments for research use, as well as provide plasma samples collected at infusion visits every 4 weeks. Participants were selected based on availability of baseline plasma p-$tau_{217}$ and clinical assessment with MMSE and DSRS.

*PET Acquisition and Processing*: Tau PET was acquired using six 5-minute frames from 75-105 minutes after injection of $^{18}$F-flortaucipir. Processed $^{18}$F-Flortaucipir PET images with uniform 6mm full-width-at-half-maximum resolution were downloaded from the ADNI archive ("Coreg, Avg, Std Img and Vox Size, Uniform Resolution"). All PET data were registered to T1-weighted images using ANTs rigid-body registration. T1-weighted images were processed as described below; an inferior cerebellar reference region was used to generate SUVR maps and mean SUVR was extracted from cortical regions after partial volume correction. Regional SUVR was used to calculate Global Tau-MaX as previously described[3]. Briefly, SUVR is converted to tau pathology index (TPI) using previously developed Gaussian-mixture models, and then thresholded to include only regions above a tau positivity threshold of 2 standard deviations above the non-pathologic distribution. TPI in tau+ regions are then multiplied by relative region size and summed to generate Global Tau-MaX, a global measure of tau burden that accounts for both magnitude and extent of disease pathology.

*MRI Acquisition and Processing*: For ADNI, participants were imaged using 3T MRI scanners to collect T1-weigthed structural images. For details of specific protocols, see www.adni-info.org. For ABC, T1-weighted images were acquired using a magnetization-prepared gradient-echo (MPRAGE) sequence [repetition time (TR) = 2400 ms, echo time (TE) = 2.24 ms, inversion time (TI) = 1060 ms, flip angle = 8°, 208 axial slices, slice thickness = 0.8 mm, field of view = 240 × 256; in plane resolution = 0.8 mm × 0.8 mm]. For both ADNI and ABC, images were pre-processed as previously described[4,5]. Briefly, images were bias-corrected and skull-stripped using ANTs prior to cerebellar, cortical, and subcortical parcellation using multi-atlas segmentation with Joint Label fusion with the MICCAI 2012 BrainColor parcellation[6–8]. ANTs cortical thickness was used to extract the mean cortical thickness from all ROIs[9]. In addition, medial temporal lobe (MTL) segmentation was performed using the T1-ASHS and CRASHS pipeline, which divides the MTL into anterior hippocampus (aH), posterior hippocampus (pH), amygdala, entorhinal cortex (ERC), Brodmann Area (BA) 35, BA36, and parahippocampal cortex (PHC), for ROI and surface-based analyses, respectively[10–13]. For ROI analysis, volume was calculated for aH, pH, and amygdala, while median thickness was extracted for cortical MTL regions.\

For MTL surface-based analyses, pointwise thickness analyses were performed using the mesh_glm tool from the CM-Rep Python package (github.com/pyushkevich/cmrep). General linear model  testing was conducted at each vertex of the MTL thickness maps and permutation testing (n = 10000) using threshold-free cluster enhancement to compute family-wise error rate-corrected p-values at each vertex.

*Plasma Collection and Analysis*: Samples were collected as previously described in ADNI[14] and ABC[15]. All samples were analyzed on a Fujirebio Lumipulse G1200 analyzer at the University of Pennsylvania ADNI Biomarker Core laboratory (n = 162) or University of Indiana (n = 363). For ADNI, p-$tau_{217}$ concentration was measured in the most recently collected sample. For ABC, p-$tau_{217}$ and $A\beta_{42}$ were measured in the most recently collected sample. For ATM, p-$tau_{217}$ was measured in the sample collected at baseline (within 4 weeks of starting anti-amyloid therapy).

*Sampled-Iterative Local Approximation (SILA)*: We generated longitudinal Tau-MaX and p-$tau_{217}$ trajectories using the silaR implementation of SILA in a combined dataset of all Aβ+ individuals in ADNI and Penn ABC with longitudinal Tau-PET and plasma data available. Briefly, SILA uses discrete sampling to determine the relationship between rate of change in a biomarker at a given biomarker level and then uses robust LOESS smoothing and Euler's method to integrate data into a single biomarker trajectory curve[16]. After defining a cutoff value for biomarker positivity, this curve can be plotted against biomarker positivity time to estimate time from biomarker positivity for a given biomarker value[16].

To determine cutoffs for Tau+, we first determined a Tau-MaX cutoff based on the 97.5%ile of CU Aβ- participants from our previously described dataset, resulting in a cutoff of Tau-MaX > 3.31[3]. We then used Receiver Operating Curve (ROC) analysis to evaluate the optimal cutoff based on Youden Index for p-$tau_{217}$ to identify those with Tau-MaX > 3.31 in this same dataset, which resulted in a cutoff of p-$tau_{217}$ > 0.3175. We used these cutoffs for SILA models of Tau-MaX and p-$tau_{217}$ trajectories, respectively, and then generated Estimated Tau

Onset Age (ETOA) for all participants using the last available observation with 3 years of interpolation and truncation of estimated ages to the ages observed in the original model. A similar approach was then used to determine Estimated Cognitive impairment Onset Age (ECOA) as described in the main methods, which was then referenced against ETOA to determine gap between ETOA and ECOA for survival analyses.

*ATM CDR-SB Crosswalk:* As CDR-SB was not available in ATM, we used a cross-walk from the Dementia Severity Rating Scale and MMSE based on historical data from Penn ADRC participants with all three measures available. This procedure followed a similar procedure as previously described for DSRS to CDR-SB[17]. The resulting formula was CDR-SB = 7.50 + 0.23*DSRS - 0.26*MMSE and had $R^2$ = 0.83.

*Sensitivity Analyses*: We repeated the primary cross-sectional analyses in ADNI treating mismatch as a continuous variable by using the residual itself as the predictor rather than mismatch group. In addition, we repeated longitudinal survival analyses in ADNI by performing separate SILA models of cognitive trajectory for each mismatch group to generate mismatch-specific ECOA in place of the single trajectory used in the primary analysis.

**eResults**

In the 258 individuals with all measures available, there was a strong association between Tau-MaX and p-$tau_{217}$ ($\beta_{std}$ = 0.63 [0.54, 0.72], $t(255)$ = 13.6, $p$ < .001), after controlling for CL (eFigure 1).

**eFigure 1.**

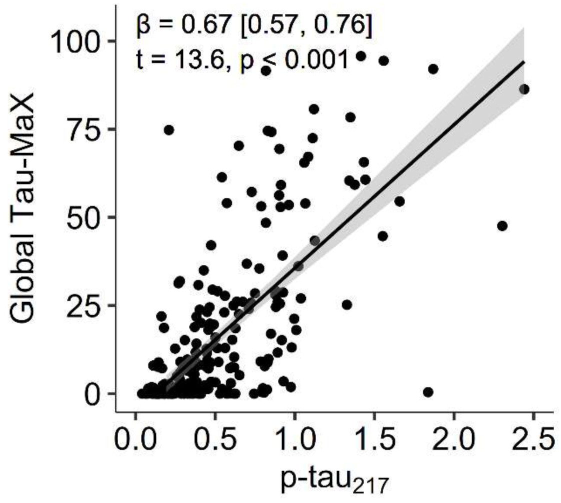


**eFigure 1. Association between p-tau$_{217}$ and Tau-MaX**. Each point represents an individual and the line shows the linear best-fit with 95% confidence of fit shown by the shaded ribbon. The *β*-coefficient [95% confidence interval], *t*-test and *p*-value for the association of p-tau$_{217}$ with Tau-MaX after controlling for Centiloids are shown.

*ROI Analyses of MTL structure in ADNI and ABC datasets*

For the ADNI tau-PET dataset, vulnerable participants showed lower aH pH, and amygdala volume ($p \leq .019$), as well as lower ERC, BA35, and PHC thickness ($p \leq .026$) compared to canonical participants (eFigure 2A). The resilient group showed the opposite pattern with higher aH, pH, and amygdala volume ($p \leq .008$), as well as greater thickness in ERC, BA35, and PHC ($p < .001$) compared to the canonical group (eFigure 2A). For the ADNI p-$tau_{217}$ dataset, vulnerable participants showed lower volume or thickness across all MTL regions ($p \leq .013$), while resilient participants showed higher volume or thickness across all MTL regions ($p \leq .009$) compared to the canonical group (eFigure 1B). For the ABC dataset, vulnerable participants showed lower volume or thickness across all MTL regions ($p \leq .007$), while the resilient group did not differ compared to the canonical group in any MTL region (eFigure 2C).

eFigure 2.

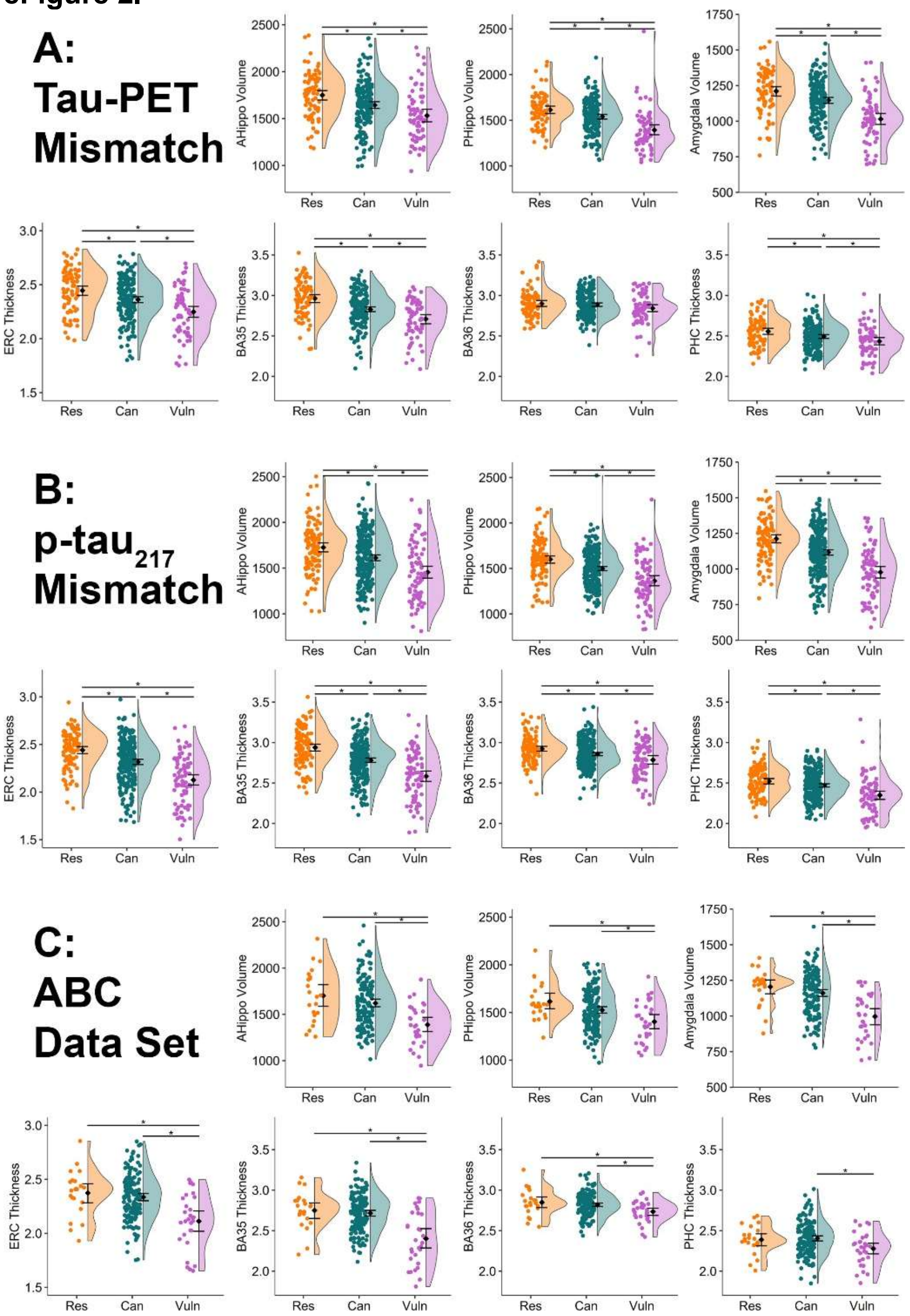


**eFigure 2. ROI results comparing mismatch groups.** Group comparisons within MTL regions in the Tau-PET **(A)**, p-tau$_{217}$ **(B)**, and Penn ABC (**C)** dataset are shown. In each panel, Anterior Hippocampus (AHippo), Posterior hippocampus (PHippo), and amygdala volume are shown in the top row and entorhinal cortex (ERC), Brodman Area (BA) 35, BA36, and parahippocampal cortex (PHC) thickness are shown in the bottom row. Each data point represents an individual and significant differences after FDR-correction are shown with bars. *$p_{FDR} < 0.05$.

*Mismatch as a Continuous Measure*

We repeated analyses treating mismatch as a continuous measure and found a negative association between mismatch residual (more positive value = more vulnerable) and volume or thickness in all MTL regions for both Tau-MaX ($p < .001$) and p-tau$_{217}$ ($p < .001$) defined mismatch after controlling for Tau-MaX (eFigure 3). We also found a negative association between mismatch residual and ERC/PHC ratio for both Tau-MaX ($p < .001$) and p-tau$_{217}$ ($p < .001$) defined mismatch (eFigure 3), as well as higher risk of αSyn+ as Tau-MaX (OR = 1.32 [1.03, 1.70], $p = .027$) or p-tau$_{217}$ (OR = 1.41 [1.15, 1.73], $p = .001$) mismatch residual increased.

**eFigure 3.**

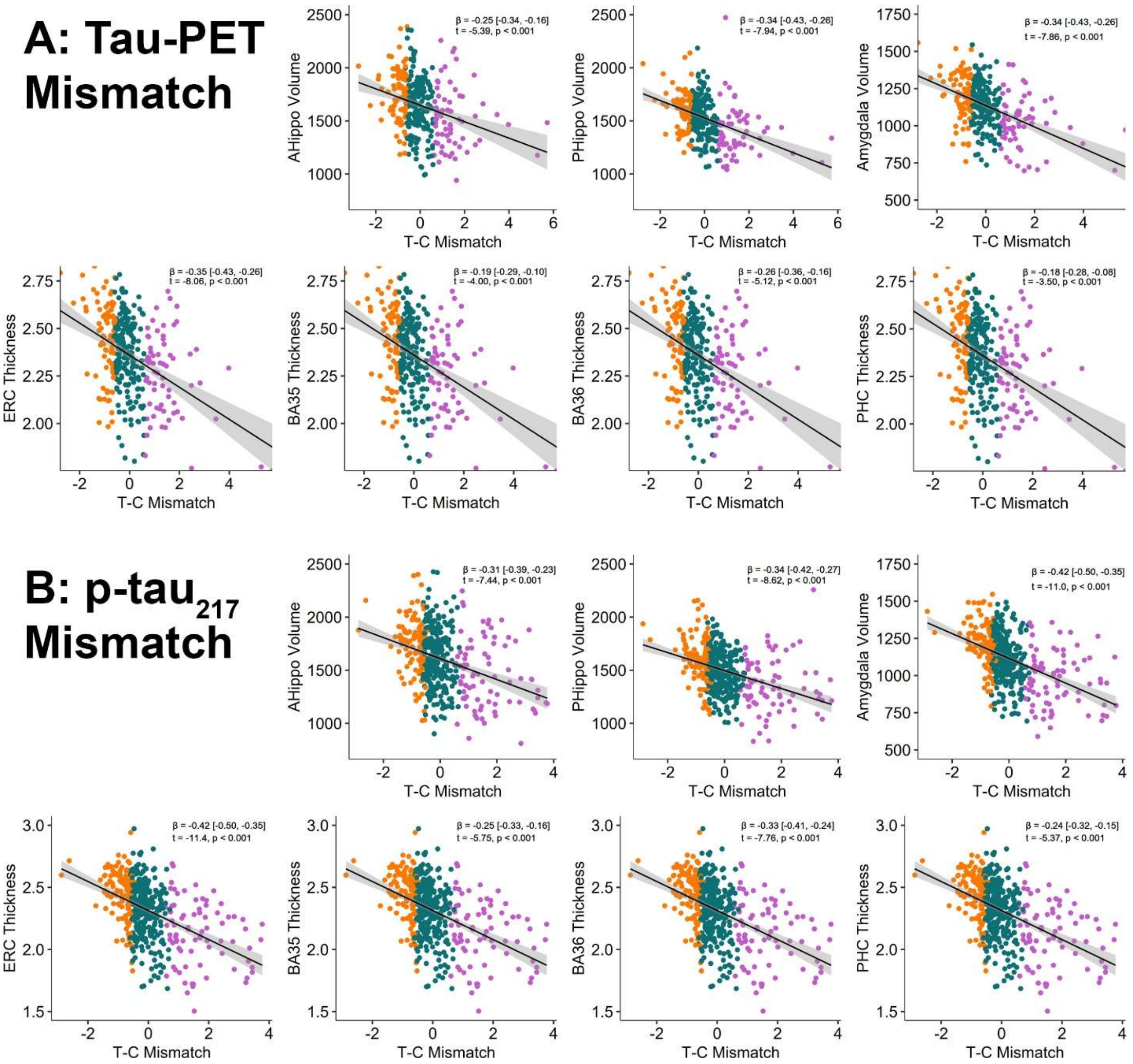


**eFigure 3. Association of continuous mismatch residual with MTL structure.** Association between the residual from the Tau-MaX **(A)** or p-tau$_{217}$ **(B)** association with CDR-SB (T-C Mismatch) and MTL structure are shown. Colors of each point represent the mismatch group assignment from group-based analyses. The standardized *β* [95% CI], *t*-test, and *p*-value for each association are shown.

*Mismatch-specific cognitive trajectories and survival*

We repeated CDR-SB SILA models using longitudinal data from each mismatch group separately to create mismatch-specific trajectories. Using this approach, we found similar results as using a common CDR-SB trajectory for all participants with vulnerable participants having higher risk for developing cognitive impairment over time (Tau-MaX: HR = 4.48 [3.38, 5.96], $p$ < .001; p-tau$_{217}$: HR = 3.04 [2.41, 3.86], $p$ < .001) and resilient participants had lower risk for developing cognitive impairment over time (Tau-MaX: HR = 0.69 [0.53, 0.88], $p$ = .004; p-tau$_{217}$: HR = 0.21 [0.16, 0.27], $p$ < .001) compared to canonical participants (eFigure 4).

**eFigure 4.**

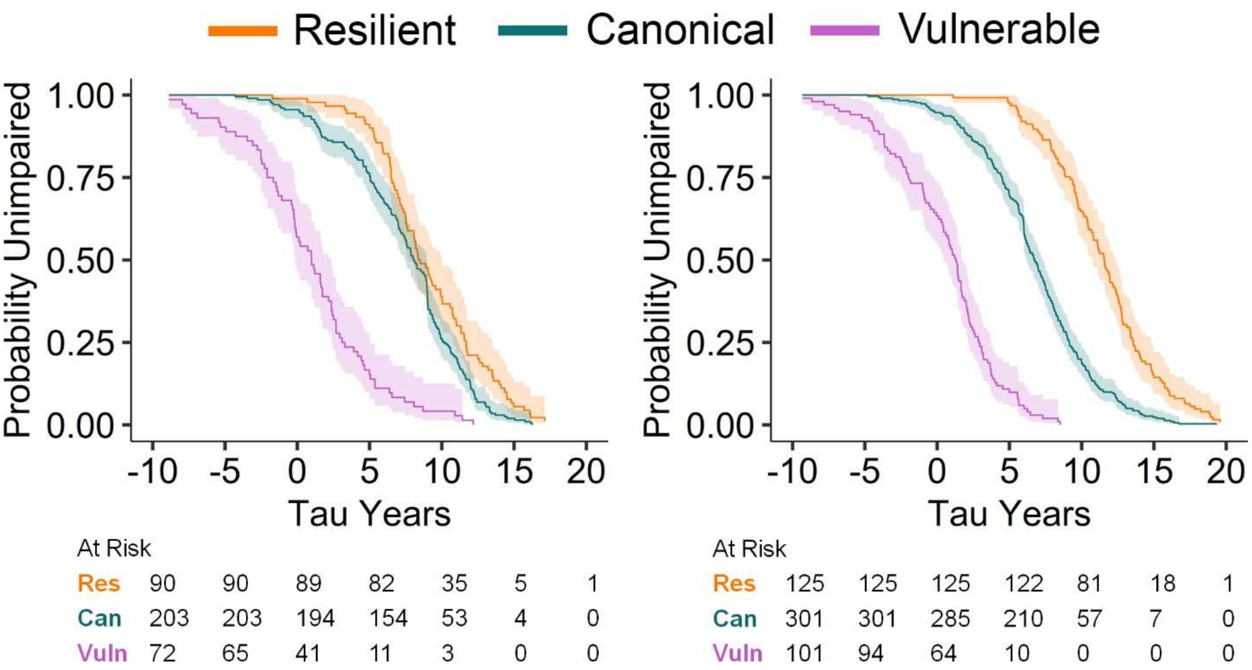


**eFigure 4. Survival using mismatch-specific SILA models for determining cognitive impairment onset.** Time surviving without clinically significant impairment (defined as CDR-SB < 4) based on years of tau positivity for Tau-MaX (left) and p-tau$_{217}$ (right) mismatch groups. Lines represent the survival curve for each mismatch group (color) with ribbon showing the 95% confidence interval of fit.

### References


1. Toledo JB, Van Deerlin VM, Lee EB, et al. A platform for discovery: The University of Pennsylvania Integrated Neurodegenerative Disease Biobank. *Alzheimer's & Dementia*. 2014;10(4):477-484.e1. doi:10.1016/J.JALZ.2013.06.003

2. Cousins KAQ, Korecka M, Yang W, et al. Comparison of plasma p-tau217 and Aβ42/Aβ40 biomarkers by race to detect Alzheimer's disease. *Alzheimer's & Dementia*. 2024;20(S8). doi:10.1002/alz.094605

3. Brown CA, Das SR, Cousins KAQ, et al. Tau Burden is Best Captured by Magnitude and Extent: Tau-MaX as a Measure of Global Tau. Published online January 14, 2025. doi:10.1101/2025.01.13.25320488

4. Das SR, Lyu X, Duong MT, et al. Tau-atrophy variability reveals phenotypic heterogeneity in Alzheimer's disease. *Ann Neurol*. 2021;90(5):751-762. doi:10.1002/ana.26233

5. Brown C, Das S, Xie L, et al. Medial temporal lobe gray matter microstructure in preclinical Alzheimer's disease. *Alzheimer's & Dementia*. 2024;20(6):4147-4158. doi:10.1002/ALZ.13832

6. Landman B, Warfield S. MICCAI 2012 workshop on multi-atlas labeling. In: *MICCAI Grand Challenge and Workshop on Multi-Atlas Labeling*. CreateSpace Independent Publishing Platform; 2012.

7. Wang H, Yushkevich PA. Multi-atlas segmentation with joint label fusion and corrective learning—an open source implementation. *Frontiers in Neuroinformatics*. 2013;7. doi:10.3389/FNINF.2013.00027

8. NJ T, BB A, PA C, et al. N4ITK: improved N3 bias correction. *IEEE transactions on medical imaging*. 2010;29(6):1310-1320. doi:10.1109/TMI.2010.2046908

9. NJ T, PA C, A K, et al. Large-scale evaluation of ANTs and FreeSurfer cortical thickness measurements. *NeuroImage*. 2014;99:166-179. doi:10.1016/J.NEUROIMAGE.2014.05.044

10. Xie L, Wisse LEM, Pluta J, et al. Automated segmentation of medial temporal lobe subregions on in vivo T1-weighted MRI in early stages of Alzheimer's disease. *Human Brain Mapping*. 2019;40(12):3431. doi:10.1002/HBM.24607

11. Yushkevich PA, Ittyerah R, Li Y, et al. Morphometry of medial temporal lobe subregions using high-resolution T2-weighted MRI in ADNI3: Why, how, and what's next? *Alzheimer's & Dementia*. Published online 2024. doi:10.1002/ALZ.14161

12. Wuestefeld A, Pichet Binette A, van Westen D, et al. Medial temporal lobe atrophy patterns in early-versus late-onset amnestic Alzheimer's disease. *Alzheimers Res Ther*. 2024;16(1):204. doi:10.1186/s13195-024-01571-z

13. Sadeghpour N, Lim SA, Wuestefeld A, et al. Developing an anatomically valid segmentation protocol for anterior regions of the medial temporal lobe for neurodegenerative diseases.

*bioRxiv*. Published online February 13, 2025:2025.02.11.637506. doi:10.1101/2025.02.11.637506

14. Schindler SE, Petersen KK, Saef B, et al. Head-to-head comparison of leading blood tests for Alzheimer's disease pathology. *Alzheimer's & Dementia*. Published online October 12, 2024. doi:10.1002/ALZ.14315

15. Cousins KAQ, Shaw LM, Chen-Plotkin A, et al. Distinguishing Frontotemporal Lobar Degeneration Tau From TDP-43 Using Plasma Biomarkers. *JAMA Neurology*. 2022;79(11):1155-1164. doi:10.1001/JAMANEUROL.2022.3265

16. Betthauser TJ, Bilgel M, Koscik RL, et al. Multi-method investigation of factors influencing amyloid onset and impairment in three cohorts. *Brain*. 2022;145(11):4065-4079. doi:10.1093/brain/awac213

17. Moelter ST, Glenn MA, Xie SX, et al. The Dementia Severity Rating Scale Predicts Clinical Dementia Rating Sum of Boxes Scores. *Alzheimer Dis Assoc Disord*. 2015;29(2):158-160. doi:10.1097/WAD.0000000000000031
